# Telephone training to improve ECG quality in remote screening for atrial fibrillation

**DOI:** 10.1101/2024.02.08.24302493

**Authors:** Kethaki Prathivadi Bhayankaram, Jonathan Mant, James Brimicombe, Andrew Dymond, Kate Williams, Peter H. Charlton, SAFER authorship group

## Abstract

**Background and Aims:** Self-recorded, single-lead ECGs are increasingly used to diagnose arrhythmias. However, they can be of variable quality, which can affect the reliability of interpretation. In this analysis of ECGs collected in atrial fibrillation screening studies, our aims were to: (i) determine the quality of ECGs when recorded unsupervised (at home); and (ii) investigate whether telephone training improved ECG quality.

**Methods:** Data was obtained from the Screening for Atrial Fibrillation to Reduce stroke (SAFER) programme, where participants recorded single-lead ECGs four times per day for three weeks using a handheld device. ECG quality was assessed using an automated algorithm, and participants who recorded >25% poor quality ECGs from days 4-10 of screening were identified for training. Telephone training to improve ECG recording technique was delivered when research team capacity permitted.

**Results:** 14,727 participants recorded 1,206,972 ECGs, of which 43,513 (3.6%) were poor quality. Most participants (51.9%) did not record any poor-quality ECGs. 1,105 (7.5%) participants met the threshold for training. Of these, 165 participants received training and 896 did not. Comparing these groups, the mean (95% confidence interval) reduction in the proportion of poor-quality ECGs per participant from before training (days 1-3) to after training (days 11-21) was 21.1 (17.5-23.5) % with training and 15.7 (14.5-16.8) % without training (p<0.05).

**Conclusion:** Most participants achieved adequate quality ECGs. For those that did not, ECG quality improved over time regardless, and training further improved ECG quality. Therefore, telephone training could be considered in atrial fibrillation screening programmes using single-lead ECG devices.

**What’s new?:**

- This is the first study to explore telephone training as a way to improve the quality of ECGs.
- We found that the quality of ECGs recorded by participants increased naturally over time, regardless of whether they received telephone training. A minimum time period of screening is required to allow for this natural improvement in quality.
- Telephone training was found to be beneficial in improving ECG quality, and was associated with greater improvement in quality than due to natural improvement alone.
- Real-time transmission of ECGs and assessment of ECG quality may be useful to identify participants who may benefit from telephone training.
- The quality of ECGs obtained at home is comparable when using an entirely remote process (postal device delivery and optional telephone training) vs. using an initial face-to-face training session (in-person device setup and training from GP practice nurses).

## Introduction

The electrocardiogram (ECG) has been used as a diagnostic investigation for cardiovascular disease and arrhythmia since its invention by Willem Einthoven in 1901 (1). Whilst the ECG was primarily developed as a diagnostic tool (2), it has also been used as an effective screening tool for cardiovascular disease so that patients who are found to have disease can be treated in the early stages before they start presenting with symptoms (3). The development of Artificial Intelligence (AI) algorithms has revolutionised ECG interpretation (4), with their use in devices such as smartwatches (5).

The quality of the ECG trace is key in successful interpretation to ensure that patients receive the correct diagnosis (6). ECG quality is particularly important when using telehealth ECG devices, where the quality of ECG recordings can be lower than that of 12-lead ECGs due to: the use of dry electrodes instead of gel electrodes; recording at the hands rather than the chest; holding the device incorrectly; and recording without clinical supervision. Three single hospital-based studies have explored the quality of single-lead ECG traces captured using telehealth devices in Belgium, Denmark and Germany (7–9) with the aim of screening for atrial fibrillation (AF). In these studies, the proportion of ECGs reported as insufficient quality for interpretation was moderately high, varying between 4% and 22% (7–9). None of these studies tested any intervention to improve ECG quality (7–9).

The Screening for Atrial Fibrillation with ECG to Reduce stroke (SAFER) programme is being undertaken in the UK to assess the effectiveness of screening for AF to prevent stroke (10). As part of this programme, participants are asked to record single-lead ECGs at home for 30 seconds four times a day. The hand-held single-lead ECG device used (Zenicor One, Zenicor Medical Systems AB, Sweden) can detect paroxysmal AF through screening, as demonstrated through a randomised controlled trial in Sweden (11). The ECG traces are analysed by the ECG Parser algorithm (Cardiolund AB, Sweden) which is able to indicate possible diagnoses such as AF (12). The algorithm is also able to detect whether an ECG trace is of adequate quality for accurate clinical interpretation or ‘poor-quality’ (i.e. the trace is of insufficient quality for accurate clinical interpretation).

One possible intervention to reduce the proportion of ‘poor-quality’ ECGs is the use of telephone calls to provide training in ECG recording technique. Telephone calls to provide training have been shown to improve adherence to a variety of interventions including exercise for cancer recovery (13) and Continuous Positive Air Pressure therapy (14). However, to our knowledge there is currently no published evidence exploring telephone calls as a possible intervention to improve self-recorded ECG quality.

Our first aim was to determine the quality of ECGs in participants who recorded them without any direct supervision (i.e. in their own homes). Our second aim was to determine whether telephone calls are effective in improving ECG quality. To investigate this, we identified participants who recorded a high proportion of poor-quality ECGs in the early stages of screening, and compared the quality of ECGs in the latter stages of screening between those who received additional telephone training and those that did not.

## Methods

### Dataset characteristics

We used data from three phases of the SAFER programme for this analysis:

1. <u>Feasibility study:</u> This phase was carried out in ten general practices from March-December 2019 (ISRCTN 16939438, REC ref 18/LO/2066). Participants aged 65 and over were invited to their general practice where a healthcare professional instructed them on how to use the handheld ECG device. Thereafter, they recorded 30-second ECG traces at home four times a day. In this phase, the duration of screening was either one, two or four weeks (different durations were used to investigate the optimal duration of screening, with three weeks being chosen for subsequent phases). Only those participants who were screened for four weeks are included in this analysis, and, to allow comparison with the other phases, data from the fourth week of screening was excluded.
2. <u>Remote feasibility study:</u> This phase was introduced as a result of the COVID-19 pandemic to assess whether screening can be carried out remotely and took place from October 2020-January 2021 (ISRCTN 72104369, REC ref 19/LO/1597). Participants aged 70 and over were recruited from three general practices and were sent an ECG device by post with written instructions on how to use it which included a link to an on-line video. Some participants received a call at the start of screening explaining how to use the device (this was determined at random to assess the value of such calls). Screening was undertaken at home by recording a 30-second ECG traces four times a day for three weeks.
3. <u>Trial:</u> Participants aged 70 and over have been randomised into the screening arm or the control arm since May 2021 (ISRCTN 72104369, REC ref 19/LO/1597). This analysis includes participants who had completed screening as of April 2023. Participants were offered a call at the start of screening to explain how to use the device, and some opted to receive this call. Those in the intervention arm were screened in the same way as for the remote feasibility study, with the same trigger for a training call.

All participants gave informed consent, and the studies were conducted in accordance with the Declaration of Helsinki, and ethical approval was given by the London Central NHS Research Ethics Committee (18/LO/2066 and 19/LO/1597).

### Training telephone calls

Training telephone calls were only implemented in the remote feasibility study and the trial. Participants who might benefit from training telephone calls were identified as follows. ECGs were transmitted from the handheld device to a server and analysed in near-real time using an automated ECG analysis algorithm (ECG Parser algorithm, Cardiolund AB, Sweden) (12). Participants who had at least 25% of their ECGs graded as poor-quality by the algorithm at any point on days 4-10 of screening were identified, and the research team made a training telephone call to these participants where possible. Whether or not a training call was received was primarily due to logistic issues such as staff capacity and participant availability when the call was placed.

During the training call, the participant was asked to describe which recording technique they were using and where possible, guidance to improve technique was sensitively provided. If appropriate and possible, during the call the participant recorded one or more further ECGs, which if graded as poor-quality by the algorithm prompted additional feedback and/or guidance. The calls carried out in the remote feasibility study and trial lasted an average (mean) of six minutes.

### Study design and data analysis

The characteristics of each dataset (i.e. feasibility study, remote feasibility and trial datasets) were reported in terms of the number of participants and ECGs and the number and proportion of participants with poor-quality ECGs at different thresholds (from 2% to 50%). Whether or not an ECG was tagged as ‘poor-quality’ was determined by the ECG Parser algorithm (Cardiolund AB, Sweden) (12). The difference between the proportion of participants with at least one poor-quality ECG and the proportion of participants with at least 25% poor-quality ECGs was examined for significance using the chi-square test in the feasibility, remote feasibility and trial datasets.

Subsequent analyses focus on those participants who had at least 25% of their ECG traces tagged as ‘poor-quality’ by the automated algorithm at any time between days 4 and 10 of screening. Since the data are non-parametric, median, inter-quartile range and range of the proportion of poor-quality ECGs per participant were calculated for days 1-3 of this sub-group, and for days 11-21. We chose these time intervals because participants who received training were intended to receive it during days 4-10 (10), and we excluded any participants who received training outside of this time period (i.e. before day 4 or after day 10 of screening). The data are presented as box and whisker plots for each of the three phases of SAFER. For the second and third phases, the data for participants that received a training call are presented separately from those that did not receive a training call (this was not applicable for the feasibility study as no participants received such telephone training). Due to the small number of participants in the remote feasibility study, for some analyses these data are combined with the trial data. The Wilcoxon signed rank test was used to identify significant differences between the proportions of poor-quality ECGs per participant between days 1-3 and days 11-21.

We also compared the differences in the proportion of poor-quality ECGs between days 1-3 and days 11-21 between participants who received the training call and those who did not. Since these differences are normally distributed, we calculated the mean difference and confidence intervals between the groups and used analysis of covariance (ANCOVA) to determine statistical significance between the intervention and control groups across both studies to ensure baseline standardisation (15). Due to the small number of participants in the remote feasibility study, for this analysis we combined these data with the trial data.

To explore the possibility of regression toward the mean (i.e. outliers early in screening tending toward the average later in screening), we also compared the differences between days 1-3 and days 11-21 in people who recorded a low proportion of poor-quality ECGs (<5%) across all three studies combined (16). Again, results were reported as median, inter-quartile range and range, and significance testing was performed using the Wilcoxon signed rank test.

Analysis was performed using Microsoft Excel and MATLAB R2023a. All results are reported to one decimal place.

## Results

### Quality of ECG recordings

Figure 1 shows examples of ECGs collected during screening. Figure 1(a) shows an ECG recorded by a Trial participant on their first day of screening, which was tagged as ‘poor quality’ by the automated algorithm. It is difficult to identify QRS complexes confidently in this ECG. Figure 1(b) shows an ECG recorded by the same participant on their final day of screening, which was not tagged as ‘poor quality’, and QRS complexes are clearly visible.

**Figure 1:**
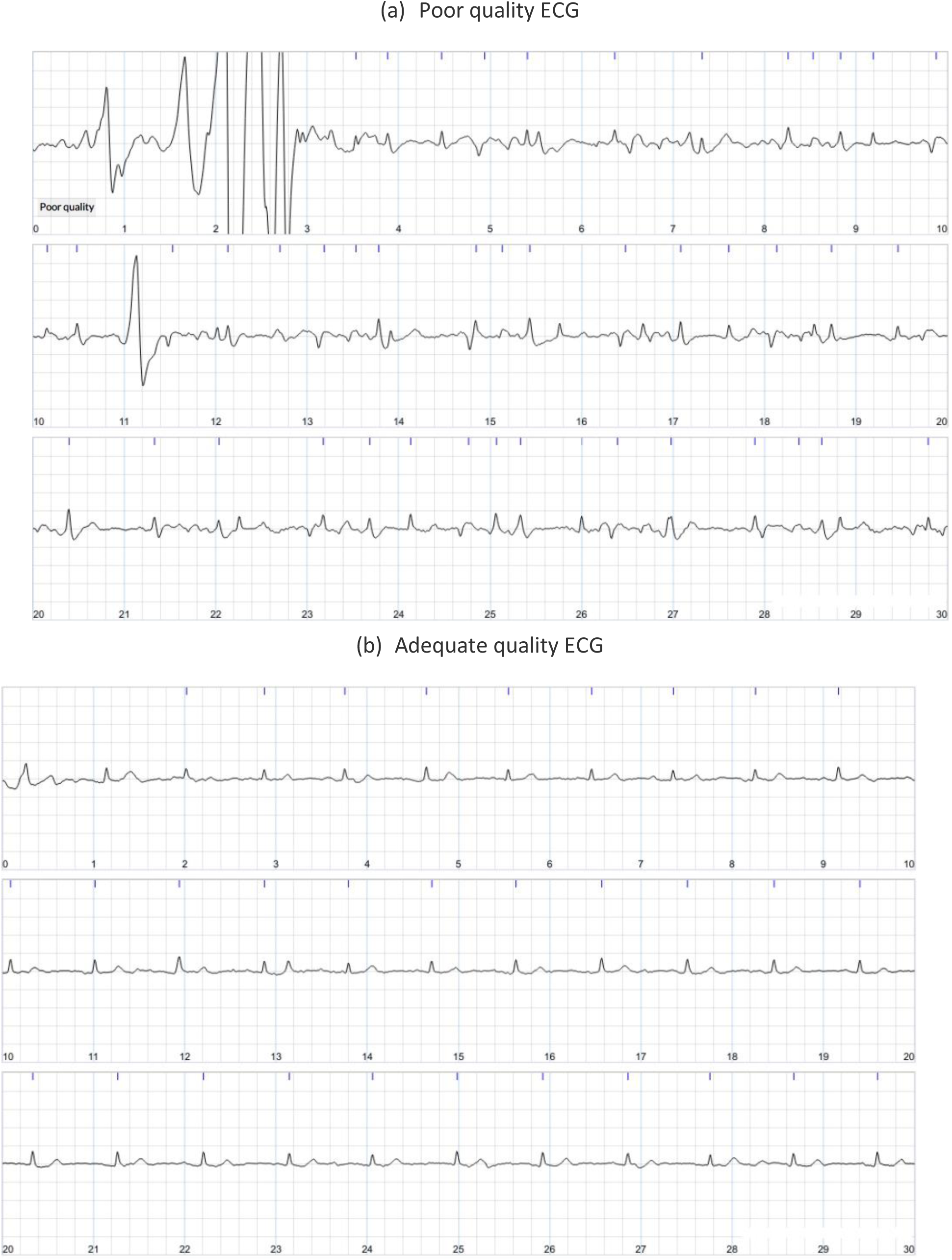
Examples of ECGs recorded during screening: (a) a ‘poor quality’ ECG recorded on day 1 of screening; (b) an adequate quality ECG recorded on the final day of screening.

Across the three phases, 14,727 participants were included in this analysis and over one million ECGs (Table 1 and Figure 2). A higher proportion of participants had at least one ECG graded as poor-quality in the trial as compared to the feasibility and remote feasibility studies respectively (48.6% versus 43.5% and 42.7%, p = 0.0353, chi-squared = 6.688 with 2 degrees of freedom). There was a tendency (non-significant) for a higher proportion of participants in the trial to have at least 25% poor-quality ECGs in days 4-10 of screening (3.4% versus 2.5% and 1.7%, p = 0.1134, chi-squared = 4.353 with 2 degrees of freedom). During the remote feasibility study, 12 (70.5%) of the 17 participants who met the threshold for a training call received one, whereas in the trial only 153 (14.7%) of the 1,044 participants eligible for a training call received one. The differences in quality of ECGs in the three phases of the study is shown in Figure 3. This demonstrates that the majority of participants had no poor-quality ECGs, approximately 30% of participants had >0% and ≤5% poor-quality ECGs, and a predominantly downward trend in the frequency of participants with an increasing proportion of poor-quality ECGs.

**Figure 2:**
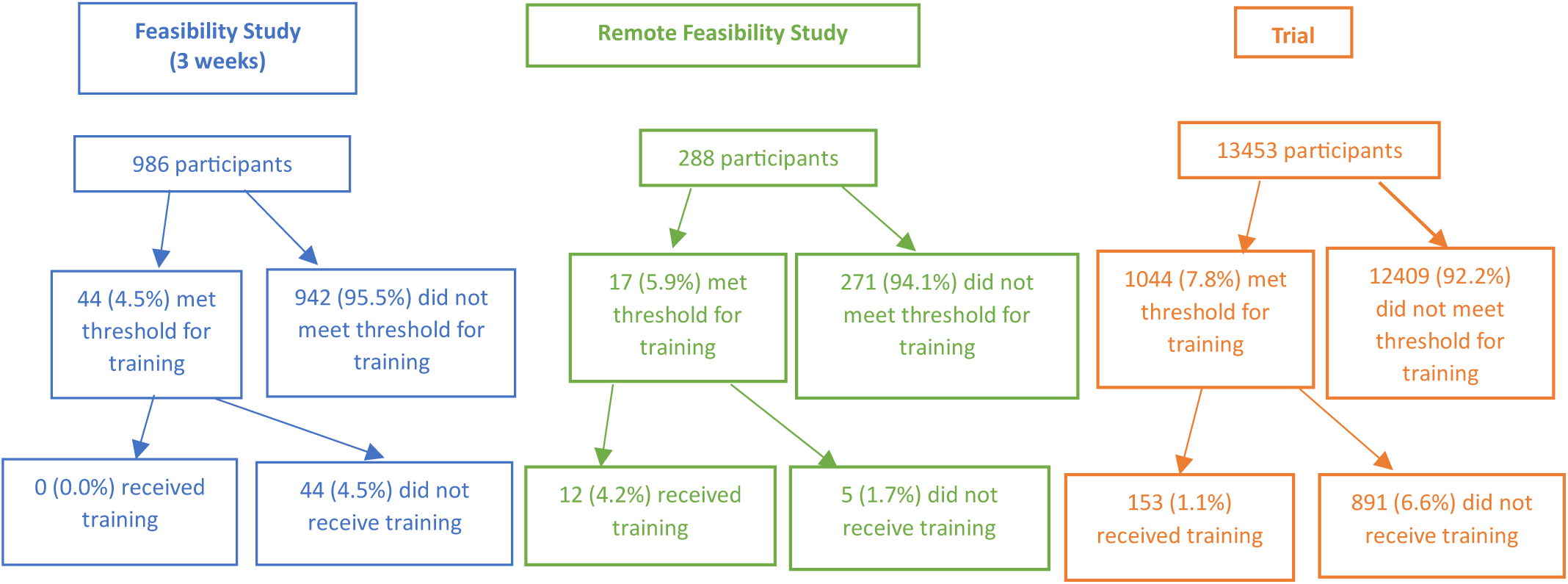
flowchart demonstrating the numbers and proportions of participants who met the threshold for a training call and subsequently either received a training call or did not receive one. The threshold for training was having at least 25% of ECGs classified as ‘poor-quality’ at any time during days 4-10 of screening.

**Figure 3:**
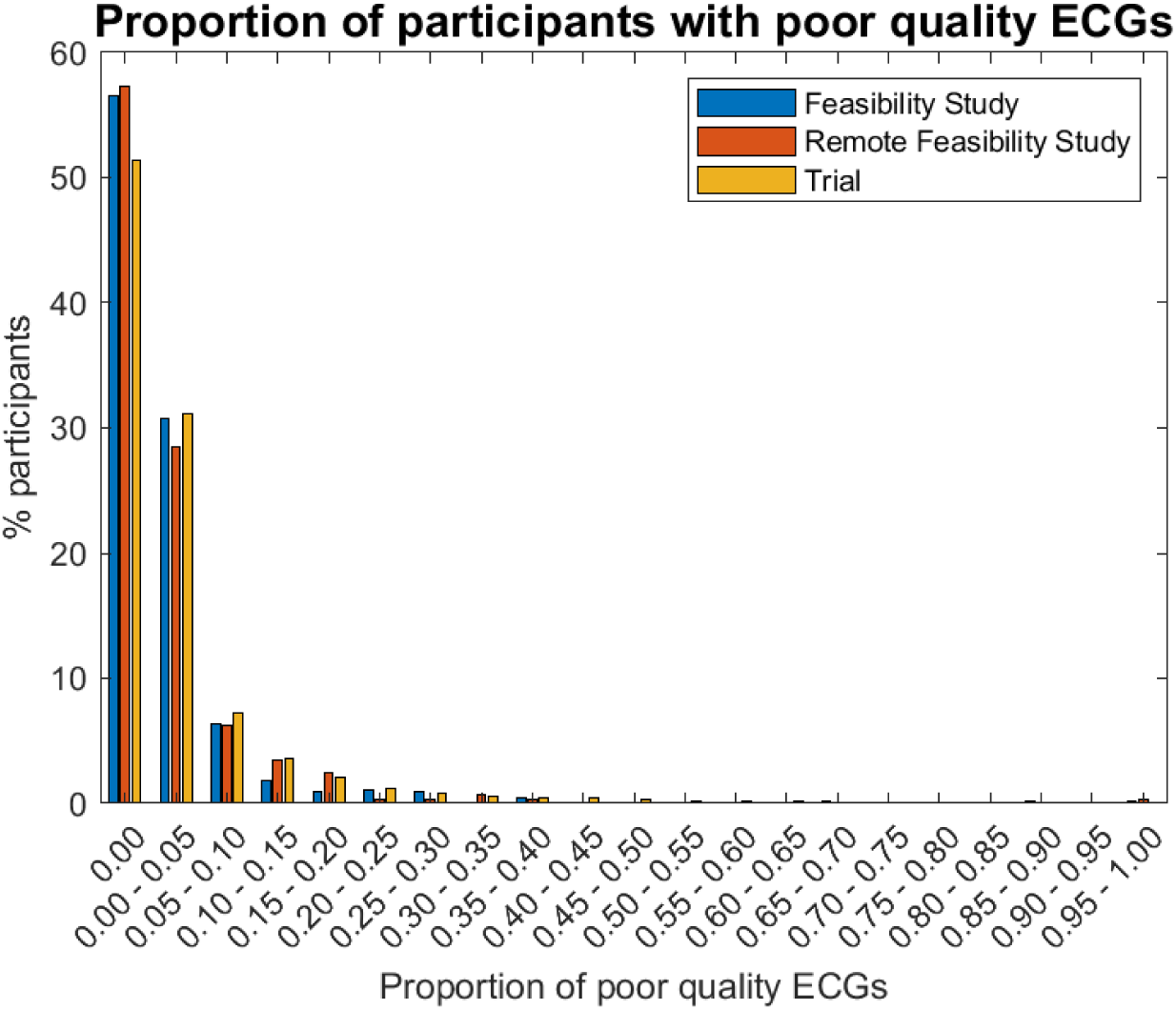
the proportions of participants with poor-quality ECGs at different thresholds in the SAFER feasibility study, SAFER remote feasibility study and SAFER trial. Note that the majority of participants did not have any poor-quality ECGs.

**Table 1:** dataset characteristics of the different phases of the SAFER programme.

|  | Feasibility Study | Remote Feasibility Study | Trial |
| --- | --- | --- | --- |
| No. participants | 986 | 288 | 13,453 |
| Age, mean (SD) | 72.7 (5.7) | 76.6 (5.9) | 76.2 (4.8) |
| Gender, no (% female) | 1,123 (52.5%) | 141 (49.0%) | 6,904 (51.3%) |
| No. GP practices | 10 | 3 | 87 |
| Total no. ECGs over 3 weeks | 79,708 | 23,259 | 1,104,005 |
| Total no. poor-quality ECGs (%) over 3 weeks | 2,225 (2.8%) | 580 (2.5%) | 40,708 (3.7%) |
| No. (%) participants<br>with at least one poor-quality ECG | 429 (43.5%) | 123 (42.7%) | 6,537 (48.6%) |
| No. (%) participants<br>with at least 25% poor-quality ECGs | 25 (2.5%) | 5 (1.7%) | 459 (3.4%) |
| No. (%) participants<br>who met the threshold<br>for training | 44 (4.5%) | 17 (5.9%) | 1044 (7.8%) |
| No. (%) participants<br>who received training | 0.0 (0.0%) | 12 (4.2%) | 153 (1.1%) |

In the feasibility study, 79,708 ECGs were recorded, of which 2,225 (2.8%) were tagged as poor-quality. 429 participants (43.5%) had at least one poor-quality ECG recording, and 25 participants (2.5%) had at least 25% of their ECGs tagged as poor-quality (Table 1). Of the 44 (4.5%) participants who met our (subsequently introduced) threshold for a training call between days 4 and 10 of screening, the proportion of poor-quality ECGs per participant reduced from a median (IQR) of 36.3 (23.5-48.5) % between days 1-3 to 20.8 (6.8-33.3) % between days 4-10 and day 21 of screening (p<0.05) (Figure 4). In the remote feasibility study, 23,259 ECGs were recorded, of which 580 (2.5%) were tagged as poor-quality. In the trial, 1,104,005 ECGs were recorded, of which 40,708 (3.7%) were tagged as poor-quality. Across all three studies, 1,206,972 ECGs were recorded, of which 43,513 (3.6%) were tagged as poor-quality.

**Figure 4:**
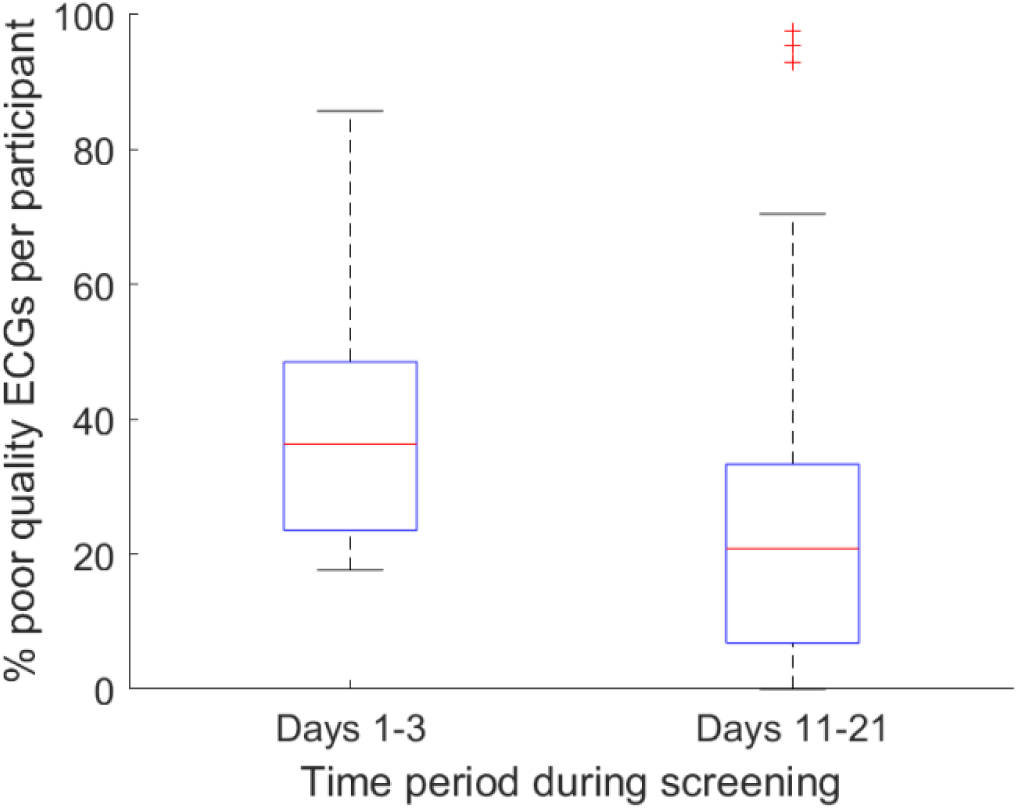
box and whisker plot demonstrating the median and quartiles of poor-quality ECGs per participant in participants who met the (subsequently introduced) threshold (>25% of their ECGs being marked as ‘poor-quality’ on days 4-10 of screening) but did not receive a training call (as there were no training calls in the feasibility study) on days 1-3 and on days 11-21 of screening in the SAFER feasibility study.

### Effectiveness of training calls

In the remote feasibility study, 17 participants (5.9%) met the threshold for training on days 4-10 of screening, of which 12 (4.2%) received training and 5 participants (1.7%) did not receive training (Figure 2). Of the participants who received training, 10 participants (3.5%) received one training call and two (0.7%) received two calls. For the participants that received telephone training, the proportion of poor-quality ECGs per participant reduced from a median (IQR) of 30.1 (21.1-45.0) % on days 1-3 to 5.2 (1.2-7.1) on days 11-21 (p<0.05) (Figure 5). The mean (95% confidence interval) difference between the proportion of poor-quality ECGs between days 1-3 and 11-21 was 25.7 (10.4-34.7) %. For the participants that did not receive training, the proportion of poor-quality ECGs per participant reduced from a median (IQR) of 25.0 (24.0-34.6) % on days 1-3 to 7.0 (3.5-25.0) % on days 11-21 (p>0.05) (Figure 5). The mean (95% confidence interval) difference between days 1-3 and 11-21 was 15.9 (-3.9-31.2) %. For the two participants that received two calls, the proportion of poor-quality ECGs after the second call increased from 18.8% before the call to 58.8% after the call for one participant and reduced from 75.0% to 2.7% for the other participant. Analysis of covariance (ANCOVA) demonstrated that for the remote feasibility study, there was no significant difference (p>0.05) in the changes in proportion of poor-quality ECGs between the participants who received telephone training and participants who met the threshold for telephone training but did not receive it.

**Figure 5:**
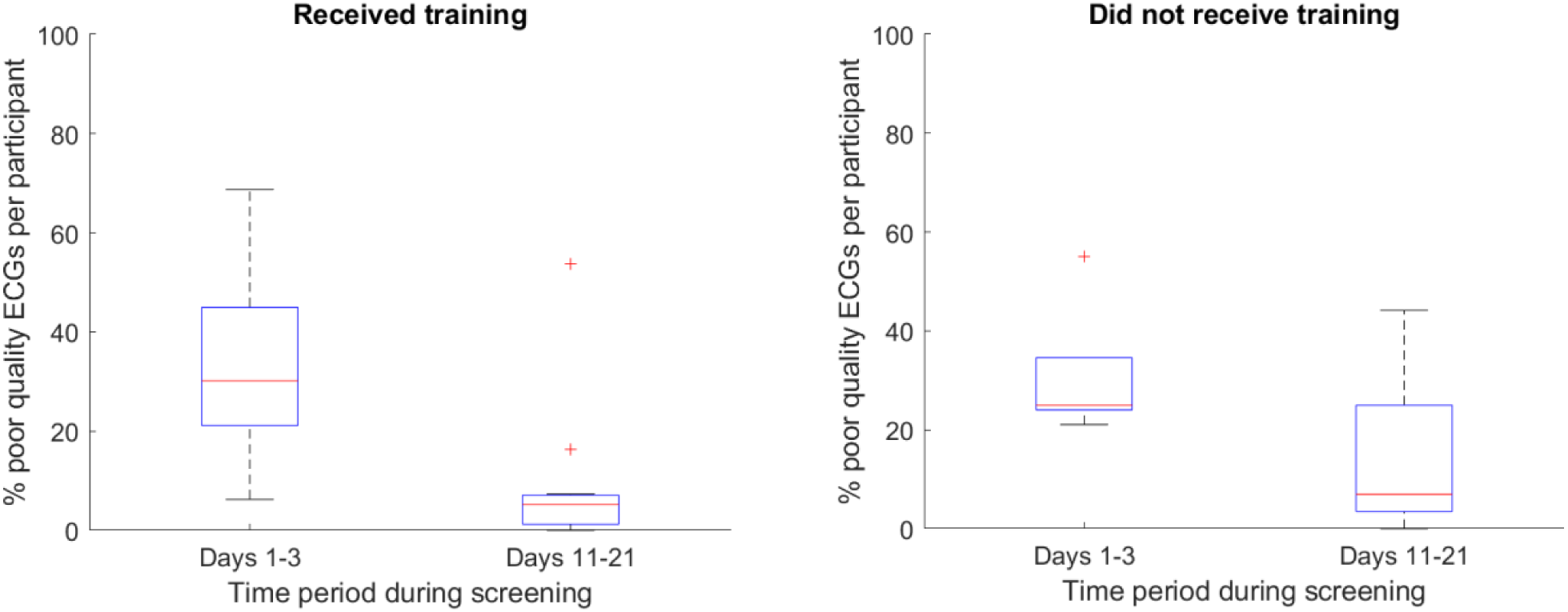
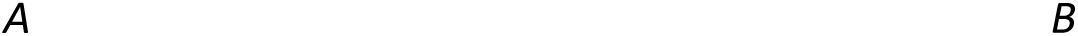
A: box and whisker plots demonstrating the median and quartiles of proportions of poor-quality ECGs per participant on days 1-3 and 11-21 of screening in participants who met the threshold (>25% of their ECGs being marked as ‘poor-quality’ on days 4-10 of screening) for a training call and received training in the SAFER remote feasibility study and B: did not receive training in the SAFER remote feasibility study.

In the trial, 1,044 participants (7.8%) met the threshold for training on days 4-10 of screening, of which 153 (1.1%) received a single training call and 891 participants (6.6%) did not receive training (Figure 2). For the participants that received telephone training, the proportion of poor-quality ECGs per participant reduced from a median (IQR) of 41.6 (31.3-50.0) % on days 1-3 to 18.6 (6.3-32.4) % on days 11-21 (p<0.05) (Figure 6). The mean (95% confidence interval) difference between the proportion of poor-quality ECGs on days 1-3 and 11-21 was 20.7 (17.2-23.0) %. For the participants that did not receive training, the proportion of poor-quality ECGs per participant reduced from a median (IQR) of 31.3 (25.0-43.8) % on days 1-3 to 14.6 (5.0-31.0) % on days 11-21 (p<0.05) (Figure 6). The mean (95% confidence interval) difference between days 1-3 and days 11-21 of screening was 15.7 (14.5-16.7) %. Analysis of covariance (ANCOVA) demonstrated that for the trial, there was no significant difference (p>0.05) in the changes in proportion of poor-quality ECGs between the participants who received telephone training and participants who met the threshold for telephone training but did not receive it.

**Figure 6:**
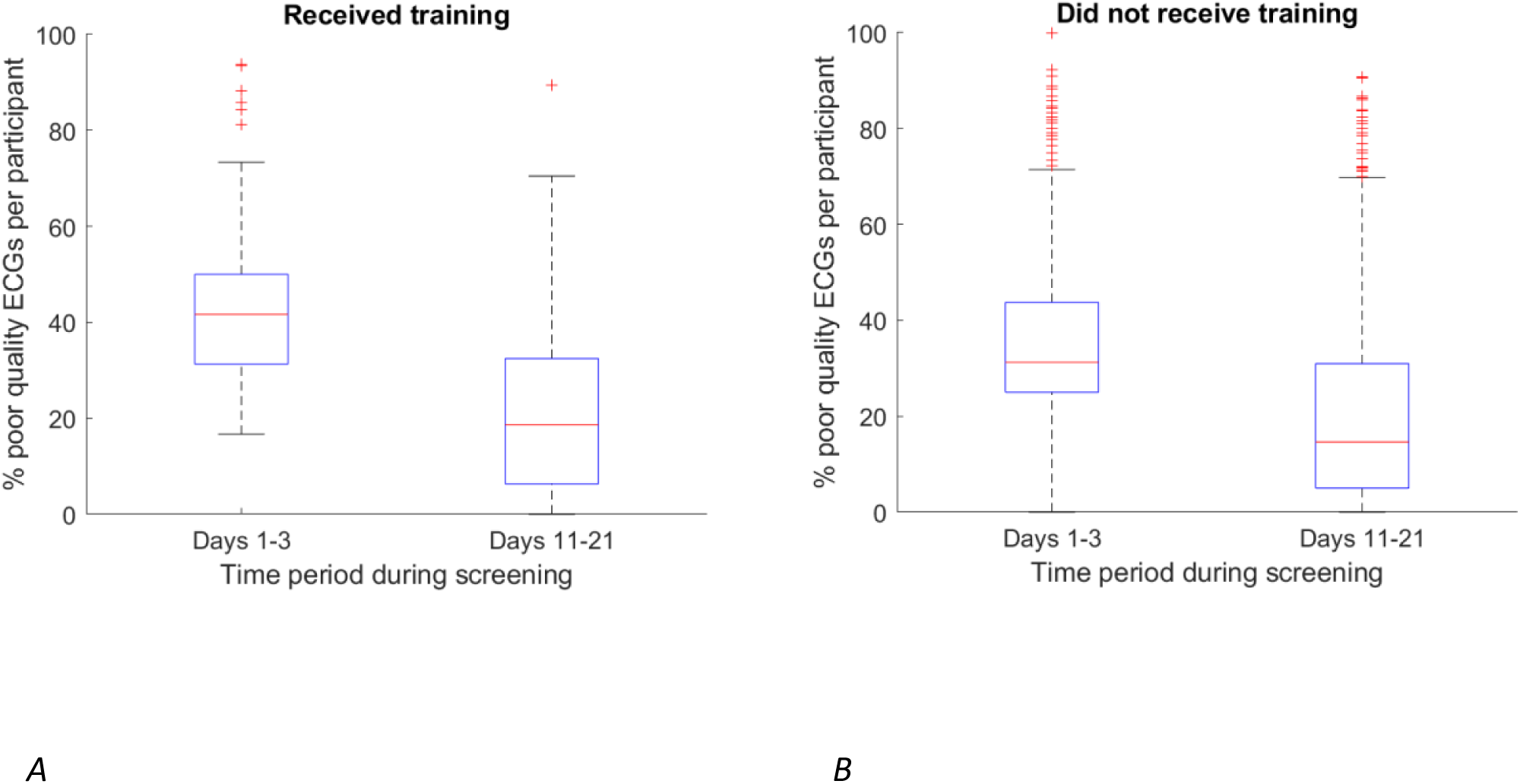
A: box and whisker plot demonstrating the median and quartiles of proportions of poor-quality ECGs per participant on days 1-3 and 11-21 of screening in participants who met the threshold (>25% of their ECGs being marked as ‘poor-quality’ on days 4-10 of screening) for a training call and received training in the SAFER trial and B: did not receive training in the SAFER trial.

Combining the results from the remote feasibility study and the trial, the mean (95% confidence interval) for the difference between the proportion of poor-quality ECGs on days 1-3 and days 11-21 was 21.1 (17.5-23.5) in the intervention group and 15.7 (14.5-16.8) % in the control group. Analysis of covariance (ANCOVA) demonstrated that there was a significant difference (p<0.05) in the changes in proportion of poor-quality ECGs between the participants who received telephone training and participants who met the threshold for telephone training but did not receive it.

Analysis of regression to the mean in participants with <5% poor-quality ECGs demonstrated that the median (IQR) proportion of poor-quality ECGs per participant was at 0 (0–0) % on days 1-3 and remained at 0 (0–0) % on days 11-21 (p>0.05).

## Discussion

### Summary of findings

Our study is the first to explore telephone training as a measure to improve the quality of ECGs in the context of screening for AF. We found that the majority of participants had ECGs of sufficiently high quality to not require further training by telephone calls, and that quality of ECGs improved over time regardless of whether or not telephone training was received. However, quality did improve further in participants with a high proportion of poor-quality ECGs who received training than in those who did not receive training. Analysis of the proportion of poor-quality ECGs across the studies demonstrated that the change to remote monitoring for the remote feasibility study did not seem to impact the proportion of poor-quality ECGs as the remote feasibility study and the in-person feasibility study (with face to face delivery) showed similar proportions of participants with different thresholds of poor-quality ECGs.

### Comparison with existing literature

We found 40-50% of participants in all three studies had at least one poor-quality ECG and 1-3% had at least 25% of their ECGs marked as poor-quality. Comparable data in the context of AF screening have not been reported in the existing literature.

In terms of the total number and proportion of poor-quality ECGs, participants across all three studies in the SAFER programme had 2-4% of their ECGs marked as poor-quality. This is similar to the geriatrics ward (3.2%) in the Belgian study where inpatients were screened for AF (7). However, the cardiology ward in the Belgian study and the German and Danish studies investigating AF screening in patients demonstrated 6.6%, 13% and 20% of their ECGs marked as poor-quality respectively (7–9). In these studies, the ECGs were all independently reviewed with either electrophysiologists (7, 9) or cardiologists (8) who manually classified the ECGs as poor-quality, unlike our study. The STROKESTOP trial used the same algorithm as our study and classified 0.99% of ECGs as poor-quality (12).

### Strengths and limitations

This is the first study to explore the use of telephone calls as an intervention to improve ECG quality. With over 14,000 participants, the study was of sufficient size to explore change over time and the impact of telephone training. Furthermore, the study included a control group, which led to the finding that ECG quality improved over time in both those who received training and those who did not. We checked that this was not explained by a tendency of outliers toward the mean (*i.e.* subjects with an outlying proportion of poor-quality ECGs early in screening tending towards the mean proportion later in screening), adding confidence to our observation that ECG quality improved over time (16).

However, allocation to the telephone intervention was not random but dependent upon team capacity. Team capacity issues are likely to explain the small decline in quality of ECGs moving from the feasibility and trial phases, when many more participants were being screened and a lower proportion of participants meeting the threshold for a training call actually received a training call. Secondly, this study was performed using only one handheld ECG device (Zenicor One) and other methods of recording single-lead ECGs are available (9). Studies using the most widely used alternative ECG device (Alivecor) have not reported ECG quality to our knowledge (17, 18).

### Implications

#### 1. Adequate quality ECGs are obtained from remote screening

We found that the majority of ECGs obtained using the Zenicor One device are of adequate quality. Furthermore, the similar results obtained in both feasibility studies shows that postal delivery of devices can lead to similar quality of ECGs as face-to-face delivery (and training) by practice nurses.

#### 2. A minimum time period of screening is required to allow for natural improvements in quality

This study has highlighted that the quality of ECGs improved over time regardless of whether or not participants received training. This is likely to reflect increasing familiarity with and confidence in using the device over time. An implication of this is that screening should be carried out over a minimum period of time in order to benefit from this natural improvement. We suggest at least two weeks of screening (preferably three) is required.

#### 3. Real time monitoring of ECG quality may be useful

This study has demonstrated that telephone training was associated with an improvement in ECG quality. This is unlikely to be due to selection bias since whether or not training was received reflected screening capacity rather than participant factors. Therefore, real-time monitoring of ECG quality may be of value. However, telephone training can be labour-intensive and there will be sufficient improvement in ECG quality in most participants without any intervention. Cost-effectiveness of real-time monitoring and telephone training needs to be assessed to determine whether it might be appropriate.

## Conclusion

Most participants of home ECG monitoring achieved sufficiently high levels of adequate quality ECGs. For the minority that did not, the ECG quality improved over time regardless of whether or not they received additional training, but additional telephone training further improved ECG quality. Therefore, such an approach should be considered if AF screening programmes using such devices are implemented.

## Data Availability

Requests for pseudonymised data should be directed to the SAFER study co-ordinator and will be considered by the investigators, in accordance with participant consent.

## Acknowledgements

This study is funded by the NIHR [Programme Grants for Applied Research Programme (RP-PG0217-20007)], and [School for Primary Care Research (SPCR-2014-10043, project 410)], and the British Heart Foundation (grant number FS/20/20/34626). The views expressed are those of the author(s) and not necessarily those of the NIHR or the Department of Health and Social Care.

## References

1. Barold SS. Willem Einthoven and the Birth of Clinical Electrocardiography a Hundred Years Ago. Card Electrophysiol Rev. 2003 Jan 1;7(1):99–104.

2. Harris PRE. The Normal Electrocardiogram: Resting 12-Lead and Electrocardiogram Monitoring in the Hospital. Crit Care Nurs Clin North Am. 2016 Sep 1;28(3):281–96.

3. Bhatia RS, Dorian P. Screening for Cardiovascular Disease Risk With Electrocardiography. JAMA Intern Med. 2018 Sep 1;178(9):1163–4.

4. Attia ZI, Harmon DM, Behr ER, Friedman PA. Application of artificial intelligence to the electrocardiogram. Eur Heart J. 2021 Sep 17;ehab649.

5. Charlton P. Peter Charlton. 2017 [cited 2023 Aug 4]. Screening for atrial fibrillation: Improving efficiency of manual review of handheld electrocardiograms. Available from: https://peterhcharlton.github.io/publication/reviewing_ecgs/

6. Zhang Y, Hou Z. An algorithm for evaluating the ECG signal quality in 12 lead ECG monitoring system. In: 2015 6th IEEE International Conference on Software Engineering and Service Science (ICSESS) [Internet]. 2015 [cited 2023 Oct 30]. p. 453–6. Available from: https://ieeexplore.ieee.org/document/7339095

7. Desteghe L, Raymaekers Z, Lutin M, Vijgen J, Dilling-Boer D, Koopman P, et al. Performance of handheld electrocardiogram devices to detect atrial fibrillation in a cardiology and geriatric ward setting. EP Eur. 2017 Jan 1;19(1):29–39.

8. Poulsen MB, Binici Z, Dominguez H, Soja AM, Kruuse C, Hornnes AH, et al. Performance of short ECG recordings twice daily to detect paroxysmal atrial fibrillation in stroke and transient ischemic attack patients. Int J Stroke. 2017 Feb 1;12(2):192–6.

9. Wegner FK, Kochhäuser S, Ellermann C, Lange PS, Frommeyer G, Leitz P, et al. Prospective blinded Evaluation of the smartphone-based AliveCor Kardia ECG monitor for Atrial Fibrillation detection: The PEAK-AF study. Eur J Intern Med. 2020 Mar 1;73:72–5.

10. Williams K, Modi RN, Dymond A, Hoare S, Powell A, Burt J, et al. Cluster randomised controlled trial of screening for atrial fibrillation in people aged 70 years and over to reduce stroke: protocol for the pilot study for the SAFER trial. BMJ Open. 2022 Sep 1;12(9):e065066.

11. Svennberg E, Friberg L, Frykman V, Al-Khalili F, Engdahl J, Rosenqvist M. Clinical outcomes in systematic screening for atrial fibrillation (STROKESTOP): a multicentre, parallel group, unmasked, randomised controlled trial. The Lancet. 2021 Oct 23;398(10310):1498–506.

12. Svennberg E, Stridh M, Engdahl J, Al-Khalili F, Friberg L, Frykman V, et al. Safe automatic one-lead electrocardiogram analysis in screening for atrial fibrillation. EP Eur. 2017 Sep 1;19(9):1449–53.

13. Mayer JE, Plumeau K. Weekly Telephone Call Impacts Outcomes of an Individualized Home Exercise Program in People Recovering From Cancer. Rehabil Oncol. 2023 Apr;41(2):89.

14. Schoch OD, Baty F, Boesch M, Benz G, Niedermann J, Brutsche MH. Telemedicine for Continuous Positive Airway Pressure in Sleep Apnea. A Randomized, Controlled Study. Ann Am Thorac Soc. 2019 Dec;16(12):1550–7.

15. Vickers AJ, Altman DG. Analysing controlled trials with baseline and follow up measurements. BMJ. 2001 Nov 10;323(7321):1123–4.

16. Bland JM, Altman DG. Statistic Notes: Regression towards the mean. BMJ. 1994 Jun 4;308(6942):1499.

17. Godin R, Yeung C, Baranchuk A, Guerra P, Healey JS. Screening for Atrial Fibrillation Using a Mobile, Single-Lead Electrocardiogram in Canadian Primary Care Clinics. Canadian Journal of Cardiology. 2019 Jul 1;35(7):840–5.

18. Ko JS, Jeong HK. Screening for Atrial Fibrillation Using a Single Lead ECG Monitoring Device. Chonnam Med J. 2021 Sep;57(3):191–6.

